# Climate Change, Place, and Mental Health in Sub-Saharan Africa: A Multi-Country Analysis of Lived Experiences Following Extreme Weather Events

**DOI:** 10.64898/2026.06.25.26356208

**Authors:** Chanelle Mulopo, Sithembiso Ndlovu, Lucy Akinyi, Alberto Muanido, Wendinmaneguede Kabre, Moussa Ouédraogo, Catija Marja Judião Maivasse, Simão Francisco José, Henry Odero, Ruth Mthembu, Luthando Zuma, Elisabeth Lindner, Marlies Craig, Nafissa Traore, Vasco FJ Cumbe, G. Nduku Wambua, Evans Omondi, Frederick Wekesah, Gillian F. Black, Collins Iwuji, Astrid Treffry-Goatley

**Author notes:** Corresponding Authors Dr. Chanelle Mulopo.

## Abstract

**Background:** Climate change is an escalating global health threat, with sub-Saharan Africa disproportionately affected due to entrenched spatial inequalities, high exposure to environmental hazards, and limited adaptive capacity. Increasingly frequent extreme weather events (EWEs), including floods and cyclones, are reshaping the material and social conditions of place, with implications for mental health and wellbeing. However, evidence remains limited, particularly multi-country qualitative research that examines how mental health impacts are produced through lived experiences of place in contexts of recurring environmental disruption and structural vulnerability. This study explored the mental health and wellbeing impacts of EWEs among individuals with lived experience of such events in Mozambique, Burkina Faso, South Africa, and Kenya, using participatory methods that centred community narratives and place-based accounts of everyday life.

**Methods:** This qualitative study employed digital storytelling as a participatory visual method to examine how EWEs are experienced and narrated across diverse socio-spatial contexts. A total of 37 participants (8–10 per country) were recruited from rural, peri-urban, and informal urban settlements with recent exposure to flooding or cyclone events. Participants produced digital stories during facilitated five-day workshops. These narratives were analysed using inductive and deductive thematic analysis informed by Braun and Clarke’s framework, with attention to the spatial and relational production of distress and coping.

**Results:** Across Mozambique, Burkina Faso, South Africa, and Kenya, findings show that the mental health impacts of EWEs are deeply embedded in place-based conditions and are cyclical, cumulative, and relational rather than confined to discrete disaster events. Participants described how repeated environmental disruptions reconfigured everyday life in place, generating ongoing uncertainty, anticipatory anxiety during rainfall periods, and acute fear during floods and cyclones. Loss of housing, livelihoods, infrastructure, and social anchors of place contributed to enduring psychological distress, which was frequently reactivated by subsequent environmental cues such as heavy rain, wind, and deteriorating physical environments. Persistent anxiety, hypervigilance, sleep disturbance, and emotional distress were reported across all sites. While social and community networks constituted critical infrastructures of care within place, these were often simultaneously overwhelmed as entire communities experienced shared disruption. Limited and delayed institutional responses further compounded spatial and social precarity.

**Conclusions:** This study provides a comparative participatory account of how EWEs shape mental health through their embeddedness in place across diverse sub-Saharan African contexts. The findings demonstrate that psychological distress is produced through the interaction of repeated environmental exposure, structural inequality, and disrupted place-based infrastructures of daily life, rather than emerging solely as a post-disaster outcome. These results underscore the need for climate-responsive mental health and psychosocial support that is integrated into place-based disaster risk governance, alongside strengthened social protection and community infrastructure that can sustain wellbeing in contexts of recurring environmental instability.

## Background

Climate change is increasingly recognised as one of the most significant global health threats of the 21st century, contributing substantially to the global burden of disease and premature mortality (Polanco Rodríguez & Álvarez Cervera, 2021; Watts et al., 2021). Beyond its physical health effects, climate change reshapes the social and environmental conditions through which health and wellbeing are produced, including the determinants of mental health. Exposure to climate-related hazards can generate psychological distress and contribute to the onset or exacerbation of mental health conditions through disrupted livelihoods, displacement, food insecurity, loss of housing and infrastructure, and the erosion or reconfiguration of social relations within place (World Health Organization, 2026). These processes highlight that the mental health consequences of climate change are not only event-based but are embedded in everyday socio-spatial conditions of life.

Although Africa has contributed only 3% of historical cumulative global greenhouse gas emissions (excluding land use) (Trisos, 2022) it is disproportionately affected by climate change due to intersecting structural and environmental vulnerabilities. These include pre-existing climatic exposure, entrenched poverty and inequality, limited adaptive capacity, and strong dependence on climate-sensitive livelihoods such as rain-fed agriculture (Ayanlade et al., 2022). Across sub-Saharan Africa (SSA), climate-related hazards including droughts, floods, cyclones, food insecurity, and water scarcity are increasingly frequent and severe. These events do not occur in isolation; rather, they interact with place-based inequalities to disrupt infrastructures of daily life, strain health and social systems, and intensify existing socioeconomic precarity (Costello et al., 2009; Sharpe & Davison, 2021; Trisos, 2022). In this sense, climate-related stressors actively reconfigure the material and social conditions of place. These impacts are further intensified by disruptions to essential infrastructures such as water, sanitation, and hygiene (WASH), which are critical to everyday health and wellbeing in many sub-Saharan African settings. Climate-related flooding and cyclones frequently damage or overwhelm WASH systems, compounding exposure to environmental contamination, displacement, and household instability (Mulopo & Chimbari, 2021; Mulopo et al., 2020). Such infrastructural breakdowns illustrate how climate-related hazards are experienced through the material conditions of place.

Evidence suggests that climate change has important implications for mental health, particularly in relation to exposure to extreme weather events (EWEs). Globally, between 25-50% of individuals exposed to EWEs experience adverse mental health outcomes, which may persist or recur over time (Barkin et al., 2021). Floods, cyclones, wildfires, and other disasters have been associated with anxiety, depression, acute stress reactions, post-traumatic stress disorder (PTSD), sleep disturbances, psychological distress, and suicidal ideation (Heanoy & Brown, 2024; Karim et al., 2024; Wambua et al., 2026). However, these outcomes are increasingly understood not only as responses to discrete events but as shaped by the ways in which repeated environmental disruptions are experienced within place. PTSD-related symptoms, including intrusive memories, hyperarousal, and avoidance behaviours, may emerge months or years after exposure and may be reactivated by ongoing environmental cues such as rainfall, wind, or deteriorating living conditions (John et al., 2007; Norris et al., 2004).

In addition to acute events, longer-term climatic changes such as rising ambient temperatures have also been associated with increased aggression, interpersonal violence, suicide, mood disorders, anxiety disorders, and higher utilisation of mental health services, including hospital admissions (Essers et al., 2025; Ryan et al., 2026). As climatic conditions intensify and EWEs become more frequent, the cumulative mental health burden is likely to increase, underscoring the importance of understanding how climate-related stressors are experienced within specific socio-spatial contexts.

Despite a growing global evidence base, important gaps remain. Much of the existing literature is derived from high-income countries and focuses on discrete post-disaster outcomes, with limited attention to how mental health impacts are produced through everyday life in contexts of chronic vulnerability and repeated climatic disruption (Alarcón Garavito et al., 2024; Brown et al., 2025; Martin et al., 2022). In particular, there is limited multi-country qualitative research from sub-Saharan Africa that examines how individuals with lived experience of EWEs understand and narrate their mental health within the context of structurally constrained and environmentally unstable places. Participatory approaches are particularly well suited to addressing this gap, as they enable the co-production of knowledge grounded in lived, situated experience.

This study addresses these gaps by examining the mental health impacts of EWEs across four SSA countries using digital storytelling, a participatory visual method that enables participants to construct and share narratives through multimedia formats (De Vecchi et al., 2016). Digital storytelling has been widely used in health research to surface marginalised voices and generate contextually grounded insights (Mpofu-Mketwa et al., 2023; Treffry-Goatley et al., 2018; West et al., 2022). In this study, it was used to capture situated accounts of floods and cyclones among individuals living in structurally vulnerable settings. By foregrounding these place-based narratives across diverse SSA contexts, the study contributes to a growing body of work in health geography and global mental health by illuminating how climate-related mental health outcomes are produced through the interaction of environmental exposure, structural inequality, and lived experience of place. The findings inform climate-responsive mental health and psychosocial support, disaster risk reduction, and adaptation strategies.

## Methods Study

### Design

This study adopted a qualitative research design using digital storytelling as a participatory visual methodology to explore the lived experiences of individuals exposed to EWEs and the associated mental health and wellbeing impacts (Mthiyane et al., 2026). Rather than treating experience as purely individual or retrospective, this approach enables the co-production of situated narratives that capture how distress, coping, and recovery are shaped through everyday life in specific socio-spatial contexts. Digital storytelling facilitates the articulation of embodied, emotional, and temporally layered experiences, supporting participants to express complex lived realities that are often difficult to capture through conventional research methods (West et al., 2022).

In addition to its narrative affordances, digital storytelling is particularly valuable in contexts marked by linguistic diversity, uneven literacy, and structural marginalisation, where conventional qualitative approaches may limit participation (Bui et al., 2026; Waluyo & Khan, 2026). Importantly, in the context of climate and health research, it enables attention to how environmental disruption is experienced and interpreted within place, foregrounding the spatial and relational dimensions of everyday life. As such, it responds to calls for more contextually grounded and epistemically inclusive approaches in climate–health research, particularly in underrepresented settings (Byskov & Hyams, 2022; Pratt, 2019).

Previous studies have demonstrated the utility of digital storytelling for examining environmental risk, disaster experiences, and community resilience in contexts affected by flooding, drought, and fire outbreaks in South Africa (Mpofu-Mketwa et al., 2023). Building on this work, the present study uses digital storytelling not only as a data collection tool, but as a means of accessing how climate-related distress and coping are produced through lived engagement with place. This approach is also consistent with broader work on knowledge translation in climate and health research, which emphasises the importance of generating contextually grounded evidence that can inform locally relevant responses in structurally constrained settings (Mulopo et al., 2025).

### Study Sites

This study was conducted across four sub-Saharan African countries: Mozambique, Burkina Faso, South Africa, and Kenya. Selected to capture diverse livelihoods, yet comparable contexts of climate-related risk where EWEs intersect with structural inequality, fragile infrastructure, and uneven adaptive capacity. Across all sites, the research focused on communities experiencing recurrent exposure to flooding, cyclones and related climatic hazards within settings characterised by socio-spatial vulnerability.

### Sofala Province, Mozambique (Beira and Dondo)

Sofala Province in central Mozambique is highly exposed to cyclones, storms, and recurrent flooding, particularly in the coastal and low-lying districts of Beira and Dondo. These areas were purposively selected due to their repeated exposure to severe climate-related disasters and ongoing processes of displacement and resettlement.

Beira City, with an estimated population of approximately 792,800 (World Population Review, 2026), is a densely populated coastal urban centre characterised by flood-prone neighbourhoods, inadequate drainage infrastructure, and high exposure to storm surges and coastal flooding (OCHA, 2019). Fieldwork was conducted in Ndunda 2, Póvoa, and Praia Nova, settlements marked by informal housing, limited basic services, and repeated disruption from extreme rainfall and flooding events.

Dondo District, located approximately 30 km inland with an estimated population of 243,723 (World Population Review, 2026), includes settlements such as Mutua and Mandruze, where many residents have been resettled following flooding in Beira and surrounding areas. These resettlement areas are characterised by constrained infrastructure development and continued vulnerability to climate variability, illustrating how displacement reshapes the spatial distribution of risk rather than resolving it.

### Kossi Province, Burkina Faso (Nouna)

The Nouna Health and Demographic Surveillance System in Kossi Province, northwestern Burkina Faso, is a long-established population surveillance site covering approximately 1,775 km² and 58 rural villages and semi-urban sectors of Nouna town. The population was estimated at 124,957 in 2019 (Barteit et al., 2023).

The region is characterised by high climate variability, including erratic rainfall, prolonged dry seasons, and extreme heat, with livelihoods predominantly dependent on subsistence agriculture. Housing is largely constructed from mud brick with corrugated iron roofing, reflecting both material constraints and heightened exposure to climate-related stressors. These conditions position the region within broader patterns of environmental precarity where climate variability directly shapes livelihood security and everyday wellbeing.

### eThekwini Municipality, South Africa (Embo, KwaZulu-Natal)

The eThekwini Metropolitan Municipality in KwaZulu-Natal has experienced increasingly frequent extreme rainfall events and associated flooding, particularly in peripheral rural and peri-urban areas. The Embo community, located approximately 38 km from central Durban, was purposively selected as a study site due to its repeated exposure to climate-related flooding within a context of limited infrastructure and service provision.

Embo is characterised by high unemployment, constrained access to healthcare services, and limited formal infrastructure, with households frequently exposed to flood damage and disrupted mobility during heavy rainfall events (Babashahi et al., 2025). These conditions reflect broader patterns of spatial inequality in the region, where peripheral communities experience disproportionate exposure to environmental risk despite proximity to a major metropolitan economy.

### Mukuru Kwa Reuben, Kenya (Nairobi)

Mukuru Kwa Reuben is a densely populated informal settlement in Nairobi characterised by overcrowding, inadequate drainage systems, and limited access to essential services. The settlement is highly susceptible to recurrent flooding during seasonal rainfall, with environmental risks exacerbated by unplanned settlement patterns and infrastructural deficits (Kering et al., 2024).

Flooding in the area routinely results in displacement, livelihood disruption, contamination of water sources, and heightened exposure to waterborne disease risks. These conditions reflect the intersection of climate variability with entrenched urban inequality, where environmental hazards are amplified by infrastructural neglect and socio-spatial marginalisation.

### Study Participants and Recruitment

Community-Based Co-Researchers (CBCRs) were recruited across the four study sites through community information sessions facilitated by public engagement and communications staff from partner research organisations with longstanding relationships in the study communities. These established relationships supported trust-building and facilitated contextually grounded recruitment processes within each site. During the information sessions, prospective CBCRs were introduced to the study aims, workshop activities, and expectations for participation.

Purposive sampling was used to recruit adults aged 18 years and older with lived experience of EWEs affecting their communities. Eligibility criteria required residence in areas that had experienced flooding, cyclones, or other severe weather-related events within the previous six years. Sampling prioritised diversity in age, gender, and socioeconomic position to capture variation in how climate-related disruptions are experienced across different social locations within place.

A total of 37 CBCRs participated across the four study sites: Kenya (n = 8), South Africa (n = 10), Burkina Faso (n = 9), and Mozambique (n = 10). Participants represented a range of livelihood and employment contexts, including subsistence farming, informal and casual labour, domestic work, community health work, and formal employment, reflecting the socioeconomic heterogeneity of climate vulnerability within each setting (Appendix Table 1).

Within the broader WEMA project, CBCRs contributed to participatory engagement, analysis, and interpretation processes. However, the present paper draws specifically on a researcher-led thematic analysis of digital storytelling narratives produced during the workshops. This analytical separation is intended to preserve methodological clarity while acknowledging that the narratives were generated within a participatory research process embedded in each site.

### Data Generation

CBCRs participated in five-day digital storytelling workshops in which they developed and shared personal narratives of their experiences of EWEs and the associated impacts on mental health and wellbeing (Table 2a and 2b). Rather than functioning solely as data collection exercises, these workshops created structured spaces for situated reflection, enabling participants to narrate how climate-related disruptions were experienced within the contexts of their everyday lives and local environments.

**Table 1a:** Activities during the five-day digital storytelling workshop.

| Day | Focus | Key Activities |
| --- | --- | --- |
| 1 | Orientation and story identification | Introductions, consent refresher, and discussion of extreme weather and mental health. Participants begin reflecting on their experiences and share initial story ideas in a facilitated story circle. |
| 2 | Story development | Participants refine their stories through discussion and feedback and begin drawing images to represent key moments in their narratives. |
| **3** | Script writing and image creation | Participants complete their story scripts and create drawings illustrating the events and emotions in their stories. |
| **4** | Image completion and digitisation | Participants finish their drawings and digitise them using tablet devices. Images are organised to match the story scripts. |
| **5** | Audio recording and digital story production | Participants record narration and combine their images and audio to produce their digital stories. The workshop concludes with a group screening and reflection. |

**Table 2b:** Prompt Research Questions Used in the DST Workshops.

| **Prompt No.** | **Prompt text** | **Purpose / focus** |
| --- | --- | --- |
| **1** | *Tell us a TRUE story about a time in the recent past (last 2–6 years) when you experienced an extreme weather event that affected your mental health.* | Elicits narrative accounts of lived experience of extreme weather events and associated mental health impacts. |
| **2** | *We would like to understand how the way you were thinking or the way you were feeling influenced your ability to get through that event.* | Explores cognitive and emotional processes shaping coping and resilience during the event. |
| **3** | *Is there anything that stands out in your memory such as something that happened or something that you or someone else did that made it more difficult or made it easier for you to bear the weather event?* | Identifies salient moments, behaviours, and social/contextual factors that facilitated or hindered coping. |

Workshops were facilitated by local social science researchers and community engagement teams in each country. The use of locally embedded facilitation teams enhanced contextual sensitivity, enabled communication in local languages, and supported culturally and socially grounded engagement throughout the storytelling process. This localisation was critical in ensuring that narratives remained anchored in participants’ lived realities within specific socio-spatial contexts.

Digital storytelling integrated narrative reflection, script development, image creation, and audio narration, allowing CBCRs to communicate their experiences through multiple expressive modalities. The process was structured to support both individual reflection and collective meaning-making through story circles and peer feedback sessions. These interactive components enabled participants to situate their experiences in relation to others within their communities, foregrounding the shared and differentiated ways in which EWEs are experienced across place.

Given the emotionally sensitive nature of the subject matter, psychosocial support was available throughout all workshops, with a psychologist, psychiatrist, or social worker present on-site. Participants were able to pause or withdraw from activities at any stage, and referral pathways for additional psychosocial support were established in all study sites to ensure ethical responsiveness to distress.

### Data Analysis

Digital stories were produced by CBCRs during the five-day workshops in the form of written and narrated scripts, which constituted the primary data for analysis. These scripts were developed through iterative reflection, peer discussion, and facilitation processes embedded within each workshop context. Community engagement teams subsequently translated the scripts into English to enable cross-site comparative analysis while preserving meaning as closely as possible to the original narratives.

All translated scripts were imported into ATLAS.ti and analysed using Braun and Clarke’s six-phase thematic analysis approach: (i) familiarisation with the data; (ii) generation of initial codes; (iii) searching for themes; (iv) reviewing themes; (v) defining and naming themes; and (vi) producing the report (Braun & Clarke, 2019). While this framework provided a structured analytic process, it was applied in a manner that prioritised interpretation of meaning as embedded in specific socio-spatial contexts rather than decontextualised thematic abstraction.

To strengthen analytic rigour across the four study sites, two digital storytelling scripts from each country were independently coded by members of the research team. Lead social scientists based in each site participated in regular weekly cross-site meetings to compare coding decisions, discuss emerging interpretations, and refine a shared coding framework. This iterative process supported the development of a coherent analytical lens while maintaining sensitivity to contextual variation across settings.

Rather than treating coding as a purely technical exercise, the analytic process was oriented towards understanding how experiences of EWEs were narrated and made meaningful within specific places. Early analysis suggested a predominantly linear temporal organisation of experiences (before, during, immediately after, and longer-term aftermath of EWEs). However, through reflexive discussion within the research team, this framing was reconsidered as it risked flattening the recurring and interconnected nature of climate-related disruptions described across sites.

The analysis was therefore reconceptualised through a cyclical and relational lens, better capturing how CBCRs described repeated exposure, anticipatory distress, and ongoing recovery as embedded within everyday life in place. This shift enabled a more spatially attuned interpretation of the data, foregrounding how mental health and wellbeing were shaped through the interaction of environmental events, infrastructural conditions, and social relations across time and place.

Cross-site analysis was used not only to ensure consistency in coding, but also to enable comparative interpretation of how similar climatic stressors were experienced differently across distinct socio-spatial contexts. This comparative approach strengthened the analytic capacity to identify both shared and context-specific processes through which EWEs are lived and narrated within place. Overall, this collaborative and iterative analytic strategy enhanced the credibility, dependability, and interpretive depth of the findings.

### Ethical considerations

Ethical approval was granted by the Biomedical Research Ethics Committee of the University of KwaZulu-Natal (Ref: BREC/00007752/2024) in South Africa; Comité d’ Ethique pour la Recherche en Santé (CERS) for Burkina Faso (reference number: 2024-12-376), AMREF Ethics and Scientific Review Committee (ESRC) for Kenya (reference number: ESRC P1792/2024), and Comité Nacional de Bioética para Saúde’s (CNBS) for Mozambique (reference number: 309/CNBS/25).

### Findings

The digital storytelling workshops conducted in Mozambique, Burkina Faso, South Africa, and Kenya explored how EWEs shaped mental health experiences within structurally vulnerable places (Figure 1). Across all four sites, participants described flooding and extreme rainfall as recurring, relational, and psychologically cumulative experiences rather than discrete disasters with clear temporal boundaries. While contexts differed across urban informal settlements, peri-urban communities, and rural agrarian places, participants’ narratives revealed interconnected and cyclical processes embedded in place, including (i) contested anticipation and environmental uncertainty in place, (ii) embodied disruption and the collapse of everyday place, (iii) layered material and relational loss across place, (iv) community-based survival and uneven recovery in place, and (v) cyclical reactivation of distress through environmental change in place.

**Figure 1.**
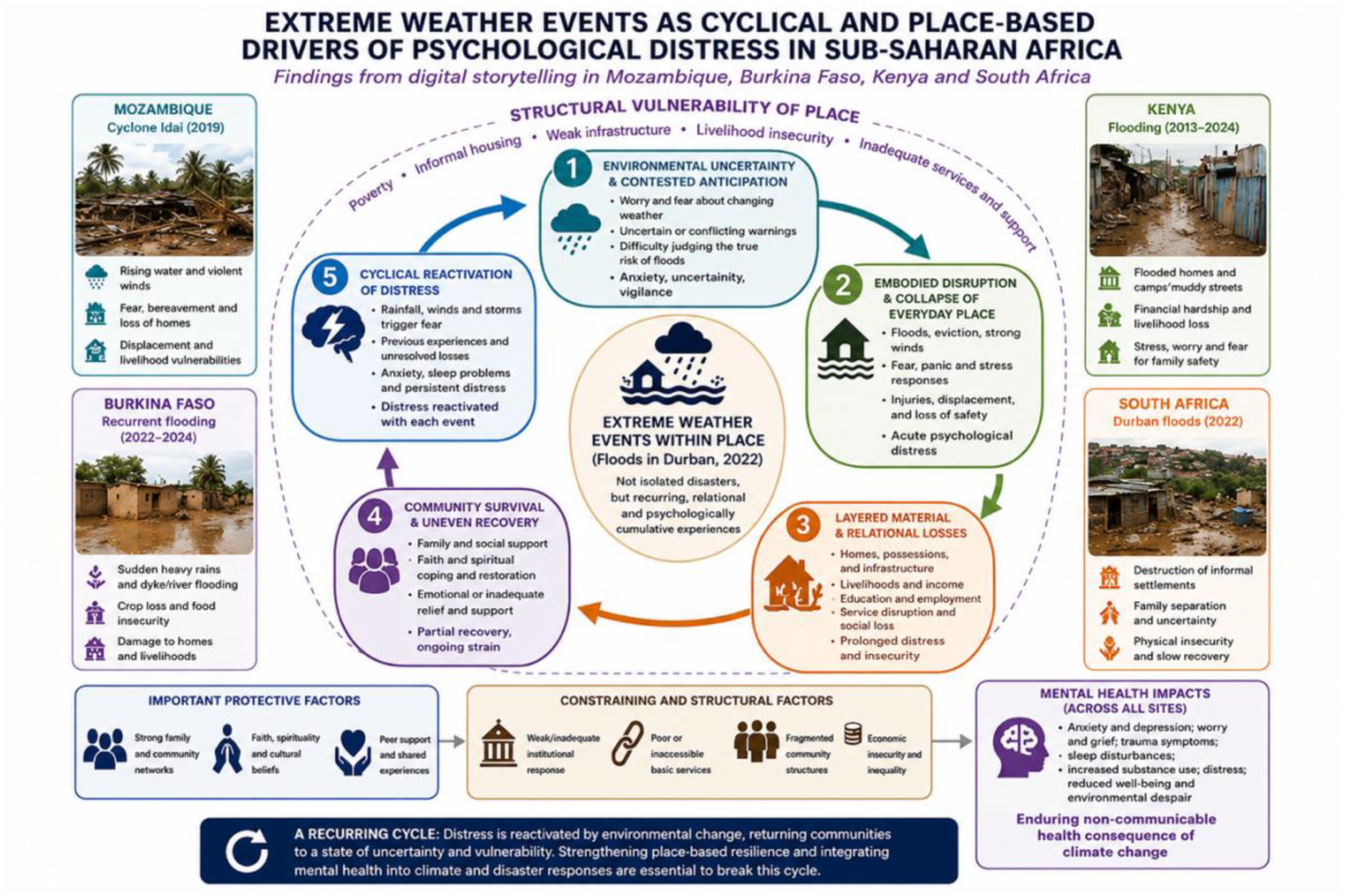
Conceptual model of the cyclical mental health impacts of extreme weather events in structurally vulnerable places across sub-Saharan Africa. (The conceptual figure was developed by the authors with assistance from ChatGPT (OpenAI) for refinement of the visual layout and wording. The authors take full responsibility for the content and interpretation presented.)

#### Contested Anticipation and Environmental Uncertainty in Place

Across all four sites, participants described living with ongoing uncertainty during periods of heavy rainfall, often attempting to interpret environmental changes, community warnings, and previous experiences to anticipate danger within place. In Mozambique, warnings regarding Cyclone Idai were disseminated through government announcements, radio broadcasts, and community leaders, yet several participants initially struggled to perceive the threat as credible. One participant recalled, *“I didn’t go to work because the government had issued a warning the day before, although I didn’t believe it*” (Soumoz, Male, 40-45 years old, Occupational Health and Safety, Mozambique). Another reflected, “*At first, I thought it was a joke*” (Luivmoz, Male, 40-45 years old, Domestic worker, Mozambique).

Similarly, participants in Burkina Faso, Kenya, and South Africa described prolonged rainfall and worsening weather conditions preceding flooding events, although many underestimated the severity of the danger until floodwaters had already entered homes or communities within place. In South Africa and Kenya, schools were dismissed early because of worsening weather conditions and flooded classrooms, reflecting institutional recognition of escalating risk within place. One participant explained, “*I was at school when the principal suddenly dismissed us because the weather was getting very bad*” (Minsou, Female, 20-25 years old, unemployed, South Africa). In Kenya, participants described receiving urgent calls from neighbours and schools warning that homes and school grounds were flooding. As one participant explained, “*I thought it was the normal rains, but it wasn’t the normal rains*”(Terke, Female, 35-39 years old, community health promoter, Kenya).

Participants in Burkina Faso similarly described repeated flooding during the rainy season between 2022 and 2024, with heavy rainfall often beginning gradually before intensifying overnight within place. Flooding was frequently experienced as sudden despite recurring seasonal exposure. One participant recalled, “*It started raining in the morning and continued until the evening*” (Perbur, Female, 35-39 years old, farmer, Burkina Faso), while another explained, “*It [started raining again] happened again around 2 a.m*.” (Yobbur, Male, 30-34 years old, farmer, Burkina Faso).

Across sites, these narratives reflected not only anticipation of environmental threat but also the normalization of recurrent flooding within places characterised by structural vulnerability, where repeated exposure complicated people’s ability to distinguish ordinary seasonal rains from life-threatening events.

#### Embodied Disruption and the Collapse of Everyday Place

Participants across all four countries described flooding and cyclones as deeply embodied and emotionally overwhelming experiences characterised by fear, confusion, panic, and immediate threats to survival within place. Participants frequently recounted waking during the night to rising water, collapsing homes, violent winds, screaming children, and darkness caused by electricity outages.

In Mozambique, participants described Cyclone Idai as unlike anything they had previously experienced within place. One participant explained, “*The wind roared from one side to the other; it was a kind of fear we had never felt before*” (Luimoz, Male, 46-50 years old, Teacher, Mozambique). In Burkina Faso, participants similarly described being abruptly awakened by children screaming as water entered their homes at night within place. One participant recounted, “*We were lying down at night when the children screamed, and we jumped up to get them*” (Konbur, Male, 40-45 years old, farmer, Burkina Faso).

In Kenya and South Africa, participants described rapidly rising floodwaters entering homes, forcing families to move belongings, rescue children, and flee in darkness within place. Participants often described intense physical exhaustion from attempting to remove water while simultaneously protecting children and elderly relatives. One Kenyan participant explained, “*We carried on trying to draw water out of the house for about six hours through the night*” (Milke, female, 30-34 years old, unemployed, Kenya). Another participant from South Africa described the panic of family separation during the floods: “*Then we shouted, ‘Mkhulu!’ because he was not in the same room as us*” (Thesou, female, 18-24 years old, unemployed, South Africa).

The emotional intensity of these experiences was amplified by the breakdown of everyday place, including darkness, cold temperatures, contaminated water, and uncertainty regarding the safety of loved ones. Participants described children crying, family members trembling, and widespread panic as homes became inundated. One participant in Burkina Faso explained, “*That day, we all cried; everyone, including the children, was in tears*” (Konbur, male, 40-45 years, farmer, Burkina Faso). Similarly, a participant from Mozambique stated, “*I was desperate, with nothing to do, feeling scared, nervous, and angry*” (Estmoz, female, 30-34 years old, domestic worker, Mozambique).

Across sites, flooding also generated acute relational trauma through injuries, deaths, near-drowning incidents, and fear of loss within place. In Mozambique, participants described searching through collapsed homes for deceased family members, especially children. One participant recounted, “*I crawled and started removing the stones, one by one. I picked up one child whose chin was completely broken*” (Soumoz, male, 40-45 years old, occupational health and safety, Mozambique). Participants in South Africa and Burkina Faso similarly described relatives becoming trapped, fainting, or being injured during evacuation attempts within place. One participant explained, “*My wife fainted, and the water’s current nearly carried her away*” (Zonbur, male, 46-50 years old, farmer, Burkina Faso).

In Kenya, participants also described fears associated with contaminated floodwaters and illness within place. One participant reflected, “*The water was full of faeces from the pit latrines*” (Terke, female, 35-39 years old, community health promoter, Kenya), while another stated, “*My house was smelly, full of mud and garbage. I was scared for my life*” (Pauke, male, 18-24 years old, causal labourer, Kenya).

#### Layered Material and Relational Loss Across Place

Across all four sites, participants described flooding as producing layered and interconnected losses affecting homes, livelihoods, relationships, health, and future stability within place. Material destruction was rarely experienced in isolation but instead triggered broader social and emotional disruptions across place.

Participants described houses collapsing, household belongings being washed away, and critical documents being destroyed within place. In South Africa, one participant explained, “*All the neighbours were outside assessing the damage. I was shocked to see that my front lawn had washed away*” (Ntosou, female, 40-45years, unemployed, South Africa). Similarly, in Burkina Faso, participants described witnessing their homes collapse while attempting to salvage belongings within place. One participant recounted, “*We returned to take a few dishes and some clothes; the house collapsed on the rest, on the chickens*”(Konbur, Male, 40-45 years, farmer, Burkina Faso).

Livelihood loss was particularly prominent in Burkina Faso and Kenya, where participants relied heavily on farming, livestock, or informal employment within place-based economies. Flooded fields, drowned livestock, and damaged homes created profound uncertainty regarding food security and survival. One participant from Burkina Faso explained, “*When we arrived, we found that the water had swallowed up our entire field*” (Perbur, female, 35-39 years older, farmer, Burkina Faso). Another participant described the emotional impact of losing crops and livestock: “*I was depressed… I wandered aimlessly, talking to myself*” (Dambur, Male, 51-55 years old, farmer, Burkina Faso).

In Kenya and Mozambique, participants similarly described financial hardship, unemployment, and difficulties meeting basic household needs following floods and cyclone impacts within place. One Kenyan participant explained, “*I did not have any money to buy clothes. It was hard for me to recover. I had no job*” (Pauke, Male, 18-24, Casual labourer, Kenya). In Mozambique, injuries sustained during Cyclone Idai prevented some participants from returning to work, contributing to prolonged economic insecurity within place.

Flooding also disrupted educational and family trajectories within place. In Burkina Faso, some participants described withdrawing children from school or considering migration because of worsening poverty and food insecurity. One participant reflected, “*The boy left the village to seek his fortune elsewhere [following the flooding], without even informing me*” (Zobbur, Male, 51-55 years old, farmer, Burkina Faso). Others described contemplating migration in search of better opportunities after losing crops and food resources. One participant explained, “*I told my wife that I would be forced to leave in search of a better life*” (Zonbur, male, 46-50 years old, farmer, Burkina Faso).

Across sites, participants described the emotional burden associated with being unable to provide for family members within place. One participant stated, “*We did our best to find food for the children so they could eat and we could have some peace*” (Konbur, male, 51-55 years old, farmer, Burkina Faso).

#### Community-Based Survival and Uneven Recovery in Place

Despite widespread destruction and distress, participants across all four sites described relying heavily on neighbours, extended family members, religious institutions, and community networks as critical infrastructures of survival within place. Community support frequently compensated for delayed, inadequate, or absent institutional assistance within place.

Participants described neighbours providing temporary shelter, helping rescue children, reinforcing damaged houses, sharing food, and offering emotional support within place. In South Africa, one participant explained, “*That same day, after our home had collapsed, we went to a neighbour’s house to find shelter*” (Silsou, female, 18-24 years old, unemployed South Africa). Similarly, in Kenya, participants described neighbours assisting children across flooded areas and landlords relocating tenants within place. One participant stated, “*My landlord moved us to one of his other houses that wasn’t flooded*” (Erike, Male, 25-29 years old, causal labourer, Kenya).

In Burkina Faso, participants described relying on neighbours and family members to stabilise damaged homes and survive financially after floods within place. One participant explained, “*I had to ask my neighbours to help me put up wooden supports to keep the house from collapsing*” (Zonbur, male, 46-50 years old, farmer, Burkina Faso). Another participant described receiving financial assistance from relatives living elsewhere: “*My daughter who lives in the city sent me a little bit of money*” (Zobbur, male, 51-55 years old, farmer, Burkina Faso).

Spirituality and faith also emerged as important coping mechanisms within place. In Mozambique, participants described praying, singing religious songs, and receiving shelter from churches during Cyclone Idai. One participant explained that her daughter began singing praise songs during the cyclone to cope with fear and uncertainty.

Although some participants received humanitarian assistance, including blankets, tents, mattresses, food, clothing, and financial support, many described institutional responses as delayed, insufficient, or unevenly distributed across place. In Mozambique, participants reported waiting years before receiving permanent housing support. One participant explained, “*After three years, the government built us a block house*” (Estmoz, female, 30-34 years old, domestic worker, Mozambique). In South Africa, participants similarly described unfulfilled promises of housing support. One participant stated, “*A local councillor came and said they would build a house for my uncle. Until now we are still waiting*” (Bonsou, female, 25-29 years old, unemployed, South Africa).

Participants also described difficult living conditions in temporary shelters and overcrowded spaces within place. In Mozambique, displaced participants reported living in camps and churches with limited privacy and safety. Some participants described additional traumatic experiences occurring within shelters, including fires and family conflict associated with prolonged displacement.

Across all sites, participants emphasized that while community support was critical, it was often insufficient to address the long-term social, economic, and emotional impacts of repeated flooding and displacement within place.

#### Cyclical Reactivation of Distress Through Environmental Change in Place

Across all four sites, participants described the psychological impacts of flooding and cyclones as ongoing and cyclical rather than confined to the immediate disaster period within place. Rainfall, thunderstorms, strong winds, and environmental changes frequently triggered fear, intrusive memories, anxiety, and emotional distress long after the events had passed.

Participants described becoming hypervigilant during periods of rain and remaining fearful that flooding would recur within place. In Mozambique, one participant explained, “*Whenever I hear that there are strong winds, I get scared and very sad*”(Marmoz, female, 18-24 years old student, Mozambique). Another participant reflected, “*Any wind, any storm that comes, I get scared, traumatized*” (Estmoz, female, 30-34 years old, domestic worker, Mozambique). Similar experiences were reported in South Africa, where participants described ongoing anxiety and fear during rainfall within place. One participant stated, “*Our lives have never been the same again because we always live in fear when it rains”* (Silsou, female, 18-24 years old, unemployed, South Africa).

In Burkina Faso and Kenya, participants similarly described rainfall as a persistent reminder of previous flooding experiences within place. One participant explained, “*Now, whenever I see threatening rain, it reminds me of what I went through in the past*” (Yobbur, Male, 30-34 years old, farmer, Burkina Faso). Participants also described difficulties sleeping, persistent rumination, stress, and emotional exhaustion within place. One participant stated, “*At times, when you lie down, you cannot sleep properly; your mind is confused*” (Konbur, male, 40-45 years old, farmer, Burkina Faso).

Participants further described prolonged sadness, hopelessness, and emotional disorientation associated with loss, uncertainty, and ongoing hardship within place. One participant from Mozambique reflected, “Idai took something very important from my life. It destroyed my life” (Luivmoz, Male, 40-45 years old, Domestic worker, Mozambique). In Kenya, participants described feeling helpless as children became sick following exposure to contaminated water and cold temperatures within place. One participant stated, “*They got sick, which made me feel frustrated because I didn’t have money for proper drugs*” (Terke, female, 35-39 years old, community health promoter, Kenya).

Across sites, these narratives demonstrate how EWEs generate enduring psychological distress that is continually reactivated through environmental conditions within place. Rather than representing isolated traumatic events, flooding and cyclones become embedded within everyday life in place as cyclical experiences of anticipation, disruption, loss, and uncertainty.

## Discussion

Across Mozambique, Burkina Faso, South Africa, and Kenya, participants’ accounts suggest that the mental health impacts of EWEs are embedded within place-based and reinforcing systems of exposure shaped by recurring environmental hazards and underlying structural vulnerabilities (Figure 2). Rather than describing floods and cyclones as discrete events with clearly bounded temporal impacts, participants portrayed distress and vulnerability as accumulating across interconnected phases of anticipation, exposure, loss, and recovery. These experiences generated cyclical feedback processes in which each episode of flooding or cyclone exposure shaped subsequent perceptions of risk, coping responses, and psychological wellbeing.

**Figure 2.**
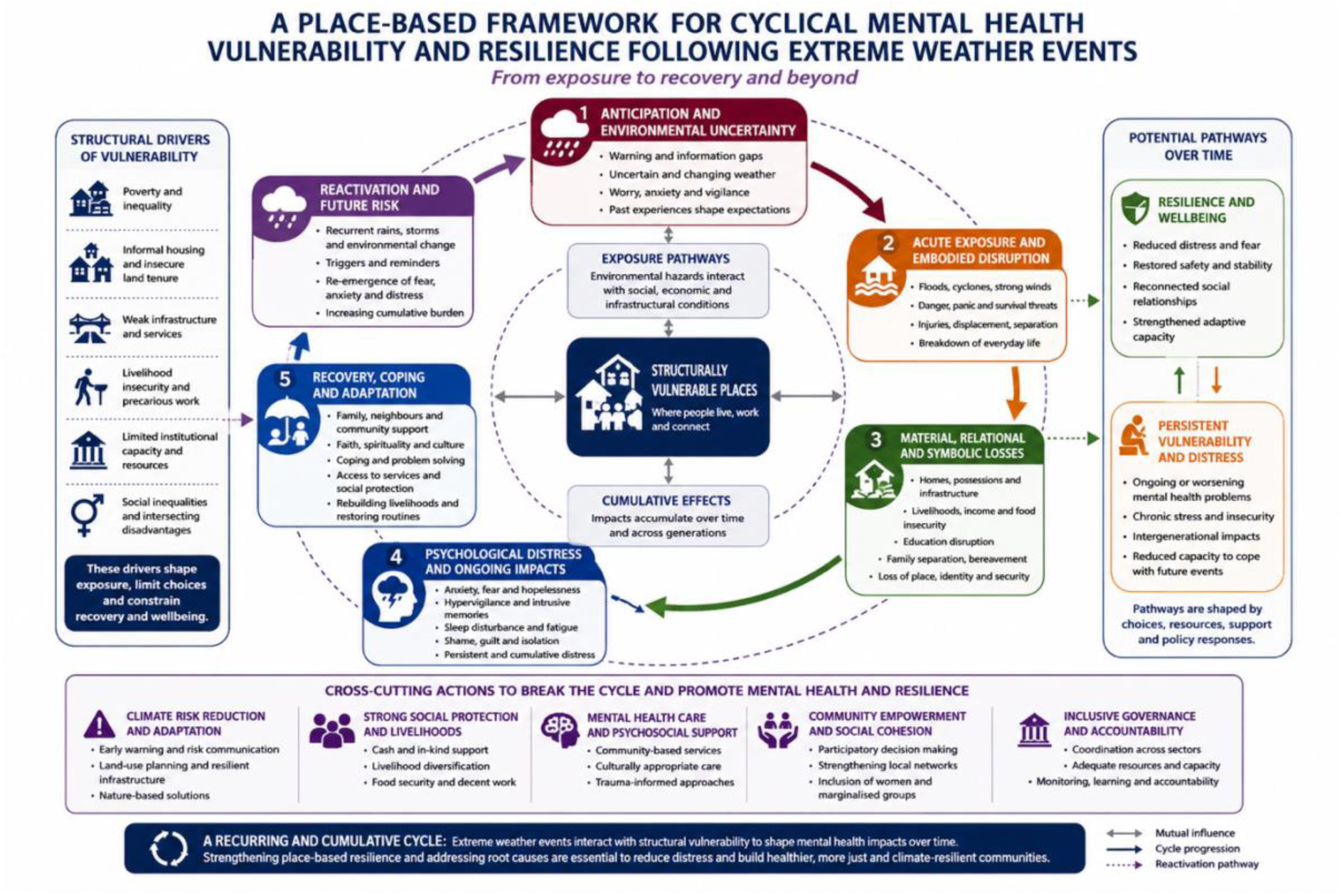
A place-based framework for cyclical mental health vulnerability and resilience following extreme weather events. The framework illustrates how floods and cyclones interact with structural vulnerabilities to shape pathways of anticipation, exposure, loss, psychological distress, and recovery, with recurrent environmental exposures reactivating vulnerability and distress over time. (The conceptual framework and figure were developed by the authors based on empirical findings from this study. ChatGPT (OpenAI, GPT-5.5) was used to assist with refining the visual layout, organisation of concepts, and wording of the figure. The authors reviewed, edited, and take full responsibility for the final content, interpretation, and presentation).

Across all four sites, participants described growing uncertainty and concern as environmental conditions worsened prior to flooding and cyclone events. During the events, this uncertainty shifted into acute fear, panic, and distress as rising floodwaters, escalating winds, destruction of homes, and threats to personal safety unfolded. These findings are consistent with Parveen and Bashir (2024) description of pre-disaster anxiety, acute crisis, and post-disaster grief associated with EWEs. However, participants’ narratives also suggest that these phases were neither discrete nor sequential. In the aftermath of floods and cyclones, participants faced extensive material, social, and emotional losses, including damage to homes, disruption of livelihoods, loss of possessions, displacement, and, in some cases, bereavement. Over time, these experiences translated into persistent emotional distress, recurring fear during periods of rainfall, helplessness, hypervigilance, and sleep disturbances. While previous research has demonstrated that flooding can result in enduring mental health consequences (Van Straten & Ncube, 2023) our findings suggest that the mental health burden associated with EWEs is continually reproduced through ongoing interactions between environmental exposure, socioeconomic precarity, and everyday life within place. Psychological distress therefore emerged not simply as a post-disaster outcome, but as part of a cyclical process through which vulnerability and mental health impacts were repeatedly reactivated over time.

The immediate vulnerability experienced by families during floods and cyclones highlights the substantial emotional and psychological burden associated with EWEs across all four study sites. Although the nature of exposure varied between contexts, participants consistently described feelings of discouragement, emotional distress, helplessness, and hopelessness following the disruption and loss of homes, livelihoods, possessions, and familiar living environments. These findings suggest that the psychological impacts of EWEs are closely tied to the disruption of place, particularly where homes and local environments function as important sources of security, identity, continuity, and social connection. This finding aligns with literature describing the emotional distress associated with the loss or transformation of environments to which people are meaningfully attached, a phenomenon conceptualised as solastalgia (Albrecht et al., 2007; Vela Sandquist et al., 2025). However, participants’ accounts also suggest that distress was not solely related to environmental change itself, but to the ways in which climate-related hazards disrupted the social and material conditions of everyday life. The loss of housing, livelihoods, and economic security often undermined participants’ capacity to provide for their families and maintain valued social roles, intensifying emotional distress. These findings are consistent with research demonstrating that climate-related disasters can exacerbate existing social and economic inequalities, with women in particular often carrying a disproportionate burden during recovery because of pre-existing poverty, caregiving responsibilities, and food insecurity (Nöthling et al., 2023). Previous studies among flood survivors in KwaZulu-Natal, South Africa, have additionally reported experiences of spiritual woundedness alongside feelings of isolation and hopelessness ((Van Straten & Ncube, 2023). While spirituality emerged primarily as a coping resource in the present study, participants similarly described profound emotional impacts associated with the disruption of everyday life and the loss of place-based sources of stability and wellbeing.

In South Africa, Kenya, and Mozambique, participants emphasised the loss and destruction of household items such as furniture, beds, utensils, and identity documents. Chaplin et al. (2020) through refugee mental health research, similarly report that the loss of personal documentation can contribute to psychological distress and insecurity. However, participants’ narratives suggest that these losses were not solely economic in nature. Household possessions, homes, and identity documents carried important symbolic and practical value because they were closely tied to feelings of security, continuity, belonging, and the ability to navigate everyday life. In the South African context, the loss of identity documents may further undermine access to social grants and other forms of government support, particularly where verification and replacement processes are lengthy and administratively complex. In Burkina Faso, although household losses were also reported, the destruction of crops and agricultural livelihoods emerged more prominently, reflecting the agrarian nature of the study setting and the central role of subsistence farming in household survival and wellbeing. These differences highlight how the consequences of EWEs are mediated by place, with the forms of loss that carry the greatest emotional and psychological significance reflecting local livelihood systems, social conditions, and material realities. Across all four sites, however, loss-whether of household assets, livelihoods, social roles, or a sense of stability and security-emerged as a central pathway through which extreme weather events contributed to psychological distress.

These findings are consistent with evidence from other climate-vulnerable settings demonstrating the close relationship between material loss and psychological wellbeing following EWEs. For example, a study conducted in informal settlements in Ghana found that flood-affected individuals experienced severe levels of depression following damage to household possessions and basic assets, with depression levels increasing significantly when household belongings were lost or destroyed (Osman et al., 2023). Similar findings have been reported in the Global North. Evidence from England suggests that the psychological consequences of flooding extend beyond immediate material losses, with individuals experiencing prolonged depression, trauma, and emotional distress following flood exposure (Waite et al., 2017). Taken together, these findings suggest that the mental health consequences of flooding are not solely attributable to hazard exposure itself, but also arise through the disruption of the material, social, and symbolic dimensions of place. Our findings extend this literature by demonstrating how these processes unfold across diverse sub-Saharan African contexts, where losses are often compounded by pre-existing socioeconomic vulnerability, limited safety nets, and repeated exposure to climate-related hazards.

In the present study, the mental health impacts of EWEs were further intensified when participants experienced the death of family members, including children, as reported in Mozambique and South Africa. These losses occurred alongside the destruction of homes, livelihoods, and community environments, creating multiple and overlapping forms of disruption. Participants’ narratives suggest that bereavement was experienced not in isolation, but within broader contexts of displacement, material loss, and uncertainty about the future. The loss of family members therefore represented both a profound personal tragedy and a disruption of the social relationships that underpin everyday life and wellbeing. Supporting this, Wambua et al. (2026) identify witnessing death and home destruction during EWEs as important risk factors for anxiety, depression, and PTSD. Our findings further suggest that the psychological consequences of EWEs are shaped by the cumulative and intersecting nature of losses, whereby bereavement, displacement, livelihood disruption, and environmental destruction occur simultaneously. Across the four study sites, these overlapping disruptions contributed to profound emotional suffering and underscore the depth and complexity of the mental health burden associated with extreme weather events.

Despite these challenges, a strong and consistent finding across Mozambique, Burkina Faso, South Africa, and Kenya was the importance of community-level social support as a critical resource for coping and recovery following EWEs. Participants described how neighbours, extended family members, religious networks, and community members assisted one another by locating missing loved ones, providing temporary shelter, sharing food and other resources, and supporting the rebuilding of damaged homes. These findings highlight the importance of social relationships and community connectedness as place-based resources that can help buffer the psychological impacts of environmental shocks. While communal coping was particularly prominent in South Africa and Kenya, similar forms of mutual assistance were evident in Mozambique and Burkina Faso despite differing social, economic, and environmental contexts.

However, participants across all four sites also highlighted an important limitation of these informal support systems. Because entire communities were often simultaneously affected by flooding and cyclones, individuals frequently had to prioritise their own immediate survival needs before assisting others. As a result, the capacity of communities to provide support was often constrained by the scale and intensity of the disaster itself. These findings suggest that while community networks constitute an important source of resilience, their effectiveness is shaped by broader structural and environmental conditions. This finding aligns with previous research demonstrating that informal support systems may become less effective during large-scale EWEs because widespread devastation forces households to focus on their own immediate survival and recovery needs (McConnell et al., 2023). Our findings therefore support growing calls for disaster response approaches that strengthen existing community capacities while recognising that community resilience alone cannot compensate for inadequate institutional support and persistent structural vulnerability.

Participants further described community support as central to their emotional recovery and ability to cope following EWEs. Neighbours, family members, religious networks, and other community actors provided not only practical assistance but also emotional reassurance during periods of uncertainty and loss. This finding aligns with existing literature demonstrating that informal social support can reduce negative cognitive and emotional outcomes following disaster exposure (Fitzgerald et al., 2020). However, participants’ accounts also highlight that emotional recovery cannot be understood solely through interpersonal support. The capacity of communities to recover was strongly influenced by broader institutional and material conditions, including access to safe housing, healthcare, financial assistance, and livelihood support. Across the study sites, delayed, inadequate, or uneven formal assistance often prolonged uncertainty and exacerbated distress. These findings suggest that mental health recovery following EWEs is shaped by the interaction between social relationships and the wider systems of support embedded within place. While community networks represent an important source of resilience, effective adaptation and recovery also require responsive disaster risk management systems capable of providing timely and equitable access to shelter, healthcare, social protection, and other essential services.

Across the study sites, participants highlighted the need for recovery strategies that extend beyond immediate disaster response and address the longer-term conditions shaping vulnerability and wellbeing. In South Africa and Mozambique, participants described returning to flood-prone areas or residing in overcrowded temporary shelters, illustrating how recovery often occurred within environments that continued to expose individuals to physical, social, and psychological risks. These conditions not only increased vulnerability to future EWEs but also contributed to ongoing concerns about safety, health, and housing security. Participants further described how the cumulative loss of homes, livelihoods, family members, and social stability resulted in prolonged economic hardship, unemployment, and uncertainty, reinforcing psychological distress long after the initial event. While these challenges were shared across sites, they also reflected place-specific vulnerabilities. In Burkina Faso, the destruction of crops and agricultural livelihoods highlighted the need for targeted agricultural support, including food assistance, access to seeds, and climate-adaptive farming strategies. In Kenya, displacement and disruptions to informal livelihoods contributed to persistent economic strain among affected households. These findings underscore the importance of place-sensitive recovery and adaptation interventions that address both immediate disaster impacts and the underlying social, economic, and environmental conditions through which vulnerability and poor mental health outcomes are reproduced over time.

The crop losses and resulting food insecurity observed in the narratives of the Burkina Faso CBCRs are consistent with broader empirical literature. Müller et al. (2023) report substantial exposure of agricultural systems to flooding, with 21% of fields affected and nearly 44% of farmers experiencing crop losses. Notably, one-third of flooded fields reportedly produced no harvest, underscoring the severity of flood impacts on food availability and household livelihoods. Beyond immediate production losses, the economic consequences of cash crop failure may extend to several months of income, contributing to cumulative processes of household impoverishment (Müller et al., 2023)

Evidence from other sub-Saharan African contexts similarly highlights how climatic stressors can disrupt agrarian livelihoods and shape mobility decisions. In Madagascar, environmental shocks such as drought and crop failure have been shown to precipitate migration among young people seeking alternative survival strategies (Hadfield et al., 2025). In the present study, participants likewise described household members relocating following agricultural losses, suggesting that livelihood disruption may function as a proximal driver of mobility in climate-affected settings.

Taken together, these findings situate climate-related shocks within a broader socio-spatial pathway linking environmental stressors, livelihood insecurity, and population movement. While migration may represent an adaptive coping strategy in contexts of acute stress, existing evidence indicates that displaced populations may also experience elevated risks of anxiety, depression, and post-traumatic stress disorder (Coşkun & Kiye, 2025; Yoo et al., 2009), highlighting the mental health consequences embedded within climate-induced mobility.

Across all four sites, participants reported enduring psychological effects, including recurrent fear during rainfall and sleep disturbances. These accounts indicate sustained distress that extends beyond the acute disaster phase and align with Bakić (2021) conceptualisation of natural disasters as acute collective events with long-term psychological sequelae. The findings also resonate with emerging scholarship on solastalgia and eco-anxiety, which foregrounds the emotional distress, anticipatory fear, and sense of insecurity associated with environmental change and recurrent climate threats (Burrows et al., 2024; Cosh et al., 2024). Taken together, these perspectives underscore that the mental health impacts of flooding and cyclones are not temporally bounded but are embedded in everyday life, shaping affective experience and perceptions of safety long after the event itself.

Importantly, the data across all sites also highlight heterogeneity in psychological response. While some individuals demonstrate adaptive coping and resilience, others experience delayed onset distress or progress to more persistent and severe mental health conditions ((Mao & Agyapong, 2021; Norris et al., 2009), reflecting uneven mental health trajectories within disaster-affected populations.

Consistent with existing literature, a substantial proportion of disaster-affected populations across diverse settings experience adverse mental health outcomes, including post-traumatic stress disorder, anxiety, and depression (Ezeonu et al., 2024; Heanoy & Brown, 2024). Taken together, these findings illustrate how EWEs operate through interconnected environmental, social, and economic pathways that shape mental health outcomes over time. These pathways are themselves embedded within broader structural conditions, including poverty, housing insecurity, fragile infrastructure, and constrained access to formal support systems, underscoring the centrality of climate justice and uneven vulnerability in shaping psychosocial risk.

In this context, the findings suggest the importance of approaches that integrate mental health support within disaster risk management systems, alongside strengthened social protection mechanisms capable of buffering households against recurrent climate-related shocks and cumulative losses in vulnerable settings.

Lastly, digital storytelling proved to be a valuable participatory approach for exploring the lived experiences of individuals affected by floods and cyclones across the four study sites. By positioning participants as co-producers of knowledge rather than passive respondents, the method enabled narratives to be shared in participants’ own words and from their own perspectives, generating rich, emotionally grounded accounts of the mental health impacts of EWEs. The collective and visual nature of the workshops further created spaces for shared reflection, validation, and connection among individuals with similar experiences.

At the same time, the process raised important ethical and methodological considerations, including the emotional difficulty of revisiting traumatic experiences, the complexities of translation across local languages and contexts, and the need for continuous psychosocial support during participatory research activities. To address this, a psychologist was present at all workshops to provide immediate support where required. Despite these challenges, the approach demonstrated considerable value for climate change and mental health research by centring community voices, enabling locally grounded understandings of distress and resilience, and supporting more inclusive forms of knowledge production.

### Limitations

This study should be interpreted in light of a few limitations. First, although it included diverse settings across Mozambique, Burkina Faso, South Africa, and Kenya, the findings are based on relatively small purposive samples drawn from structurally vulnerable communities affected by flooding and cyclones. As such, the results reflect context-specific lived experiences and cannot be considered generalisable to all populations affected by EWEs across sub-Saharan Africa.

Second, the study relied on retrospective accounts of traumatic events, which may be subject to recall bias and influenced by the emotional intensity of the experiences being narrated. Third, the use of multiple local languages and translation across sites may have introduced subtle shifts in meaning, despite efforts to preserve participants’ intended narratives during transcription and analysis. In addition, while digital storytelling enabled rich and emotionally grounded accounts, the group-based setting may have influenced disclosure, with participants potentially selectively sharing experiences they felt comfortable articulating in a collective space.

## Recommendations for Future Research

Despite these limitations, the study offers important comparative insights into the cyclical and cumulative mental health impacts of EWEs across diverse African contexts. Future research would benefit from longitudinal designs that examine how psychological distress unfolds over time following recurrent climate-related disasters, particularly in settings exposed to repeated shocks.

There is also a need for research focusing on specific vulnerable groups, including children, older adults, people with disabilities, and individuals with pre-existing mental health conditions, to better understand differentiated risk and recovery trajectories. Further studies should examine protective factors that support resilience at household and community levels, including the role of social protection systems, social networks, and culturally grounded coping strategies.

In addition, future intervention research should evaluate integrated approaches that combine disaster risk reduction with mental health and psychosocial support within resource-constrained settings. Finally, participatory methods such as digital storytelling warrant continued use and refinement in climate and mental health research, particularly for their capacity to centre community voices, generate locally grounded understandings of distress and resilience, and inform more contextually responsive policy and practice.

## Author approval

All authors have seen and approved the manuscript.

## Data availability statement

All data produced in the present study are available upon reasonable request to the authors

## Acknowledgements

We would like to acknowledge our funder, the Wellcome Trust, for the funding received to conduct the study. We sincerely acknowledge the CBCRs for their dedication and participation in the study.

## Appendices

| **CBCR Code** | **Site** | **Story Title** | **Occupation** | **Gender** | **Age** |
| --- | --- | --- | --- | --- | --- |
| Helmoz | Mozambique | Fury of Idai | Domestic worker | F | 35-39 |
| Luimoz | Mozambique | Destruction of Idai | Teacher | M | 46-50 |
| Soumoz | Mozambique | Sleepwalkers of Cyclone Idai | Occupational Health and Safety (OHS) | M | 40-45 |
| Luimoz | Mozambique | Marks of Idai | Domestic worker | F | 25-29 |
| Luivmoz | Mozambique | Idai Took My Son | Domestic worker | M | 40-45 |
| Hilmoz | Mozambique | Idai Left a Sign | Driver | M | 46-50 |
| Animoz | Mozambique | The Tragic Death of My Son | Domestic Worker | F | 30-34 |
| Delmoz | Mozambique | Cyclone Idai Destroyed My Life | Domestic worker | F | 45-50 |
| Estmoz | Mozambique | Memories of Cyclone Idai | Domestic worker | F | 30-34 |
| Marmoz | Mozambique | The Fury of Idai, Beira in Screams and Tears | Student | F | 18-22 |
| Zobbur | Burkina Faso | After the Flood, Hope | Subsistence farmer | M | 51-55 |
| Yobbur | Burkina Faso | Inner Suffering | Subsistence farmer | M | 30-34 |
| Zonbur | Burkina Faso | In the Face of Adversity | Subsistence farmer | M | 46-50 |
| Dakbur | Burkina Faso | To Cry in the Rain | Subsistence farmer | F | 46-50 |
| Perbur | Burkina Faso | Everything was Washed Away | Subsistence farmer | F | 35-39 |
| Konbur | Burkina Faso | The Distress | Subsistence farmer | M | 40-45 |
| Dambur | Burkina Faso | Struggle for Survival | Subsistence farmer | M | 51-55 |
| Dembur | Burkina Faso | A Destructive Rain | Subsistence farmer | M | 30-34 |
| Konebur | Burkina Faso | The Sorrow caused by the Flood | Subsistence farmer | M | 46-50 |
| Ntosou | South Africa | The fence | unemployed | F | 30-34 |
| Ntosou | South Africa | United | unemployed | F | 40-45 |
| Nonsou | South Africa | My wardrobe | unemployed | F | 46-50 |
| Minsou | South Africa | My books | unemployed | F | 18-24 |
| Bonsou | South Africa | My life change | unemployed | F | 25-29 |
| Lutsou | South Africa | The blue file | unemployed | M | 18-24 |
| Thesou | South Africa | Life changing | unemployed | F | 18-24 |
| Sbosou | South Africa | Never the same | unemployed | M | 18-24 |
| Balsou | South Africa | My home | unemployed | F | 46-50 |
| Silsou | South Africa | The difficulty of my life | unemployed | F | 18-24 |
| Janke | Kenya | Unforgettable Demolition | Unemployed | Female | 18-24 |
| Erike | Kenya | A Flooded Shanty | Casual labourer | Male | 25-29 |
| Milke | Kenya | Outrageous Flooding | Unemployed | Female | 30-34 |
| Ashke | Kenya | For My Mom | Casual labourer | Male | 18-24 |
| Terke | Kenya | Flooded Shack | Community health promoter | Female | 35-39 |
| Roske | Kenya | Ubomozi wa Mvua | Casual labourer | Female | 35-39 |
| Pauke | Kenya | I almost Lost My Life | Casual labourer | Male | 18-24 |
| Zubke | Kenya | My Kids | House help | Female | 30-34 |

The names in this table and the manuscripts are Pseudonyms

